# Annual risk of hepatitis E virus infection and seroreversion: insights from a serological cohort in Sitakunda, Bangladesh

**DOI:** 10.1101/2023.10.28.23297541

**Authors:** Amy Dighe, Ashraful Islam Khan, Taufiqur Rahman Bhuiyan, Md Taufiqul Islam, Zahid Hasan Khan, Ishtiakul Islam Khan, Juan Dent Hulse, Shakeel Ahmed, Mamunur Rashid, Md Zakir Hossain, Rumana Rashid, Sonia Hegde, Emily S Gurley, Firdausi Qadri, Andrew S Azman

**Author notes:** Corresponding author, Johns Hopkins Bloomberg School of Public Health, 615 N Wolfe St, Baltimore, MD 21205. Equal contributions (Emily S Gurley, Firdausi Qadri, Andrew S Azman).

## Abstract

Hepatitis E virus (HEV) is a major cause of acute jaundice in South Asia. Gaps in our understanding of transmission are driven by non-specific symptoms and scarcity of diagnostics, impeding rational control strategies. In this context, serological data can provide important proxy measures of infection. We enrolled a population-representative serological cohort of 2337 individuals in Sitakunda, Bangladesh. We estimated the annual risks of HEV infection and seroreversion both using serostatus changes between paired serum samples collected 9 months apart, and by fitting catalytic models to the age-stratified cross-sectional seroprevalence. At baseline, 15% (95CI: 14-17%) of people were seropositive, with seroprevalence highest in the relatively urban south. During the study, 27 individuals seroreverted (annual seroreversion risk: 15%, 95CI: 10-21%), and 38 seroconverted (annual infection risk: 3%, 95CI: 2-5%). Relying on cross-sectional seroprevalence data alone, and ignoring seroreversion, underestimated the annual infection risk fivefold (0.6%, 95CrI: 0.5-0.6%). When we accounted for the observed seroreversion in a reversible catalytic model, infection risk was more consistent with measured seroincidence. Our results quantify HEV infection risk in Sitakunda and highlight the importance of accounting for seroreversion when estimating infection incidence from cross-sectional seroprevalence data.

## Introduction

Hepatitis E virus (HEV) is an important cause of acute jaundice in populations with limited access to safe drinking water across South Asia and Africa [1–5]. Transmitted from person-to-person via faecal-contaminated water, HEV genotypes 1 and 2 (HEV-1 and 2) carry a high risk of death if contracted during pregnancy [6] and regularly cause outbreaks, with particularly long-lasting transmission documented in displaced persons camps [7]. In Bangladesh, HEV is the leading cause of hospital-attended acute jaundice [3]. Although more recent data are lacking, an analysis of verbal-autopsies from 1998-2007 estimated that 19-25% of maternal deaths were associated with jaundice, suggesting that HEV may be a key cause of maternal mortality nationally [2].

The risk factors driving HEV infection are not fully understood and attempts to slow transmission through emergency water and sanitation interventions have had limited success [8]. In the absence of effective treatment, vaccination is a promising tool to avert cases and deaths. Whilst an efficacious vaccine exists [9], the lack of reliable burden estimates is one of several barriers preventing the World Health Organisation (WHO) from recommending its routine use [10,11]. Sparse surveillance data and differing model assumptions mean that estimates of morbidity and mortality attributable to HEV vary widely, rendering them difficult to interpret [4,5,12,13]. Understanding the drivers of HEV infection and improving incidence estimates can greatly support both global and local decision makers.

Although not a direct measure of disease incidence, serological data can provide important proxy measures of infection. Age-stratified cross-sectional seroprevalence is often used to estimate the rate at which seronegative individuals become infected with a pathogen [14] and has previously been used to estimate HEV infection incidence though past approaches have had several limitations [4,5]. Despite evidence that antibodies to HEV wane over time [15,16], models have ignored seroreversion, which cannot always be reliably estimated from cross-sectional seroprevalence [17]. Additionally, such approaches have assumed infection risk does not vary with age or time, which may not hold for HEV. Collecting serum samples from the same individuals at different time-points can overcome some of these limitations with observed serostatus changes providing a direct measure of seroincidence and seroreversion.

In this study, we aimed to fill several gaps in our understanding of HEV infections by enrolling a population-representative longitudinal serological cohort of 580 households in an HEV-1 endemic region in Bangladesh [18,19] to explore infection risk factors and estimate the annual risk of infection, and the rate of seroreversion. As a secondary goal we aimed to compare the concordance of estimates of infection incidence derived from cross-sectional data to those observed in the longitudinal cohort.

## Methods

### Serosurvey design

We tested serum samples from a population-representative cohort recruited between March 2021 and February 2022 with the original aim of estimating *Vibrio cholerae* O1 seroincidence in the Sitakunda sub-district of Chattogram, Bangladesh (Figure S1). Households were recruited through a previously described two-stage sampling process [20]. Briefly, we first divided Sitakunda into 1km^2^ grid cells and randomly sampled cells weighted by the number of household structures identified by satellite imagery. We then randomly sampled structures to visit within each grid cell weighted by whether they were single- or multi-story units. For each household, we sought consent from the household head and attempted to enrol all members ≥1 years old. Study staff administered a questionnaire covering household-level infrastructure, assets and sanitation facilities to household representatives, and an individual-level questionnaire on demographics and drinking-water sources to all consenting household members. In addition, ∼5ml of venous blood was collected from each consenting household member (∼3ml for those <5yrs). Enrolled households were visited at a subsequent timepoint approximately 9 (range 7-11) months from baseline, to ask follow-up questions and repeat blood collection.

### Sample testing

Paired serum samples from the two survey rounds were tested for anti-HEV immunoglobulin (IgG) at icddr,b using commercially available Wantai HEV IgG ELISA kits (Wantai Biological, China). Following the manufacturer’s instructions, samples with a standardized optical density >1.1 were considered seropositive, those <0.9 were considered negative and those between 0.9-1.1 borderline. Borderline results were excluded from analyses.

### Statistical analysis

#### Seroprevalence and risk factor analyses

We estimated baseline seroprevalence and 95% confidence intervals, accounting for household sampling survey design, using the Rao-Scott method, implemented in the R package “survey” [21]. We created smoothed maps of household seroprevalence using inverse distance weighting and assessed the spatial autocorrelation by estimating the semivariogram for household seroprevalence. We explored the relationship between individual- and household-level variables and baseline seropositivity using mixed effect logistic regression models to account for household-level random effects. Firstly, we estimated univariate odds ratios of seropositivity for variables pertaining to demography, drinking-water and sanitation, history of jaundice and livestock keeping. We then constructed a multivariable model including those variables that were statistically significant (p ≤ 0.05) in univariate analyses and those identified *a priori* as potentially causally related to HEV exposures.

Drinking-water sources and sanitation facilities were categorized as improved and unimproved based on definitions from the WHO/UNICEF Joint Monitoring Programme (JMP) service ladders for drinking water [22] and sanitation facilities [23].

#### Annual risks of infection and seroreversion

We estimated the annual risk of infection of seronegative individuals (from here on referred to as just the annual risk of infection) and seroreversion of seropositive individuals using two methods: (1) using the observed changes in seroprevalence between study visits, and (2) fitting catalytic models to age-stratified seroprevalence data from a single study visit.

The annual risk of infection was first estimated by dividing the number of individuals who seroconverted over the course of the study period, *n*_*sc*_, (i.e., baseline seronegative individuals who became seropositive) by the total number of person-time at risk during the study period (total person-time for those who remained seronegative throughout, *pt*_*sn*_, plus half the person-time for those who seroconverted, *pt*_*sc*_) as described by equation (1). Similarly, the annual risk of seroreversion was first estimated by dividing the number of baseline seropositive individuals who became seronegative, *n*_*sr*_, by the total person-time at risk (total person-time for those who remained seropositive throughout *pt*_*sp*_, plus half the person-time for those who seroreverted, *pt*_*sr*_) as described by equation (2). We assumed on average individuals seroconverted or seroreverted at the midpoint of their time in the study.

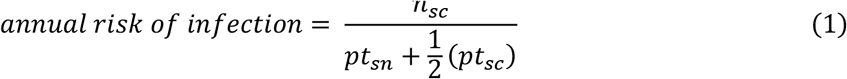

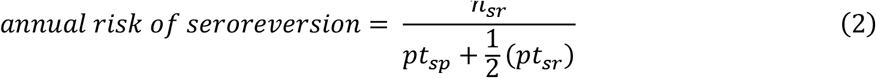

Alternative estimates of the annual risk of infection (often presented as the rate known as the force of infection when estimated in this way) were produced by fitting two catalytic models of seroconversion to the cross-sectional age-stratified seroprevalence data collected at baseline. In Model 1 we assumed no antibody waning, which has been the traditional approach to estimating HEV infection incidence [4,5]. In Model 2, due to evidence of seroreversion in our empirical data and in previous studies [15,16], we allowed for seroreversion at rate, ρ. Since previous work has shown that the annual risk of infection and the seroreversion rate often cannot be reliably estimated from cross-sectional seroprevalence data simultaneously due to identifiability issues [17], we used our empirical estimates for the rate of seroreversion in Model 2. For both models, we initially considered infection risk to be constant across age and time, then repeated model fitting whilst allowing the risk to vary by age group as has been done previously [24,25]. Model solutions are presented in the Supplementary Material. The seroprevalence data was assumed to be binomially distributed. Models were fitted within a Bayesian framework using a Hamiltonian Monte Carlo algorithm implemented within rstan [26,27]. We used a uniform prior between 0 and 1 for the per capita annual risk of infection, and between 0 and 10 for the per capita annual risk of seroreversion to conservatively include previous measures of HEV antibody persistence which have ranged from months to many years [15,16,28,29]. Model fit was assessed using Leave One Out Cross-Validation (LOO-CV) as implemented in the loo R package [30].

To estimate the annual number of HEV infections we multiplied the annual risk of infection, based on observed seroconversions within the cohort, by the estimated population of Sitakunda in 2021. We extrapolated the age-stratified population counts from the 2011 National Census-[31], assuming that the population grew by 1.5% each year between 2011 and 2021. Since we did not include <1-year olds in our survey we subtracted 20% of the population count for 0-4 year olds based on the age distribution presented in the US Census Bureau International Database for Bangladesh in 2021 [32].

### Ethical review

Adult study participants provided written, informed consent. Parents or guardians of all participants <18 years were asked to provide consent on their behalf, with those 11-17 years old also providing written assent. The protocols for the original study and extension to test samples for HEV antibodies, were approved by the icddr,b research and ethics review committee and the Johns Hopkins Bloomberg School of Public Health institutional review board.

## Results

### HEV seroprevalence at baseline

Between 27 March and 13 June 2021, 2337 individuals from 580 households were recruited to the serological cohort, and 2301 (98%) were tested for anti-HEV IgG antibodies (Table S1). At enrolment, 15% (95% confidence interval [CI]: 14-17%, ICC: 0.05, design effect: 1.15) of the sampled population of Sitakunda had antibodies indicating past HEV infection (which we refer to as being “seropositive”). Seroprevalence was significantly higher in males (20%, 95%CI: 17-22%) than females (12%, 95%CI: 10-14%) (Figure 1A), with the difference becoming more apparent during adulthood (Figure 1B). Seroprevalence was low in children and increased until approximately age 40 before plateauing (Figure 1B). Household seroprevalence was higher in populations living in the relatively urbanized south, near Chattogram city, with one notable cluster in the southeast (Figure 1C). The seroprevalence in this cluster was 46% (95%CI: 38-54%), more than 3-fold higher than the average. Compared to just 4% of the overall sample, 20% of individuals within this cluster reported that their primary drinking water source had been unavailable at least once in the month prior to the baseline survey (Table S1). When comparing demographic characteristics, a larger proportion of individuals in the cluster were male (53% compared to 46%), and more households had a monthly income <10,000TK, shared sanitation facilities, and kept livestock (Table S1). While we did detect some areas with elevated seroprevalence, across the Sitakunda, we did not detect strong spatial correlation in household seroprevalence (Figure S2).

**Figure 1.**
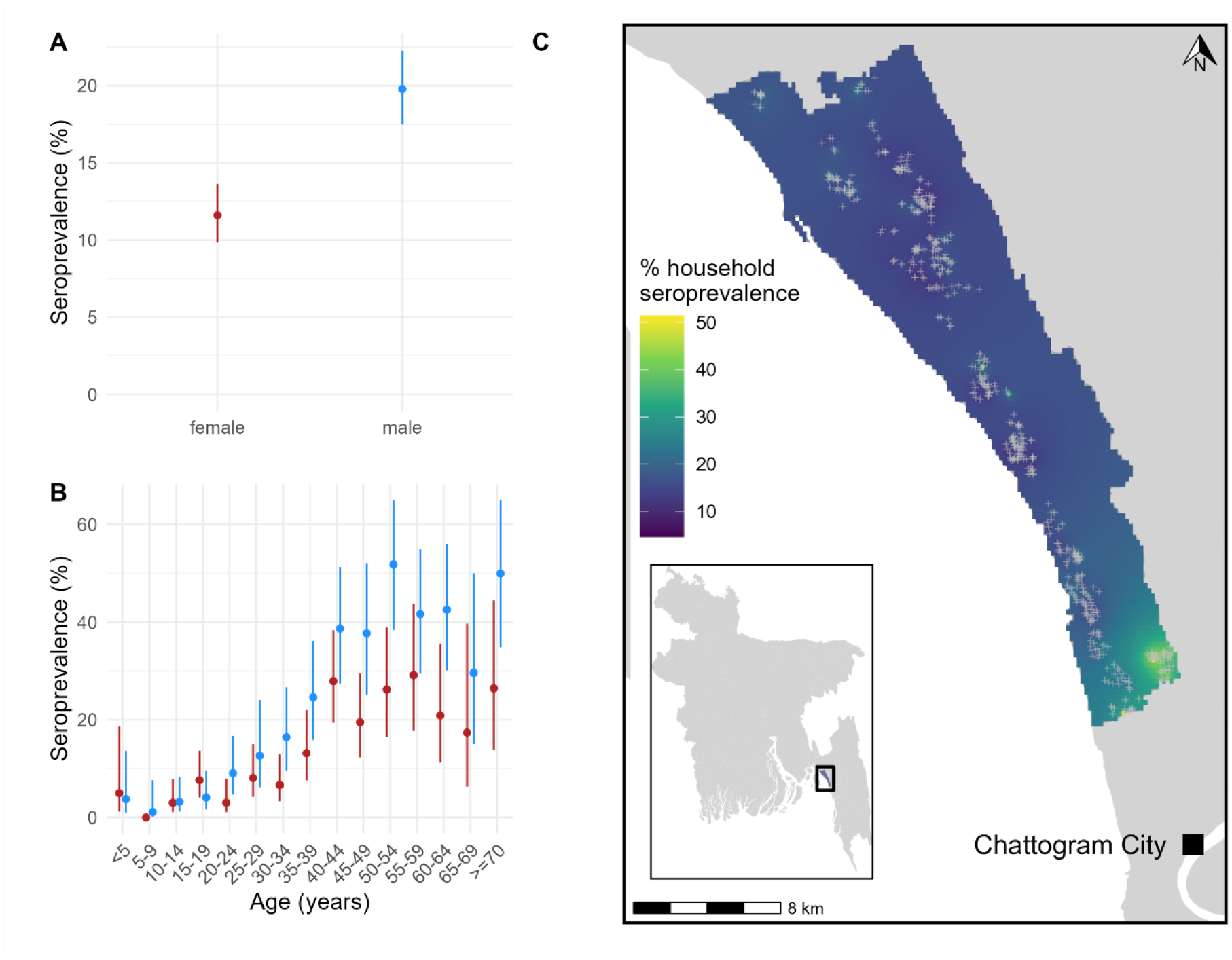
**A**. Baseline seroprevalence by sex, with 95% confidence intervals. **B**. Baseline seroprevalence stratified by age and sex, with 95% confidence intervals. **C**. Smoothed spatial variation in baseline household seroprevalence across Sitakunda. Grey crosses represent the location of sampled households.

To further understand the sex differences in seroprevalence and to identify other potential individual- and household-level risk factors for seropositivity, we conducted logistic mixed effects regression analyses. In multivariable models, we found significantly increased odds of seropositivity among those ≥40 years, those reporting that their primary water source was unavailable at least once in the past month and those with business or other occupations outside the home (Table 1). Including both sex and occupation in the multivariable model attenuated the effect size for sex (1.4, 95% CI 1.0-2.0) to the limit of statistical significance.

**Table 1.**
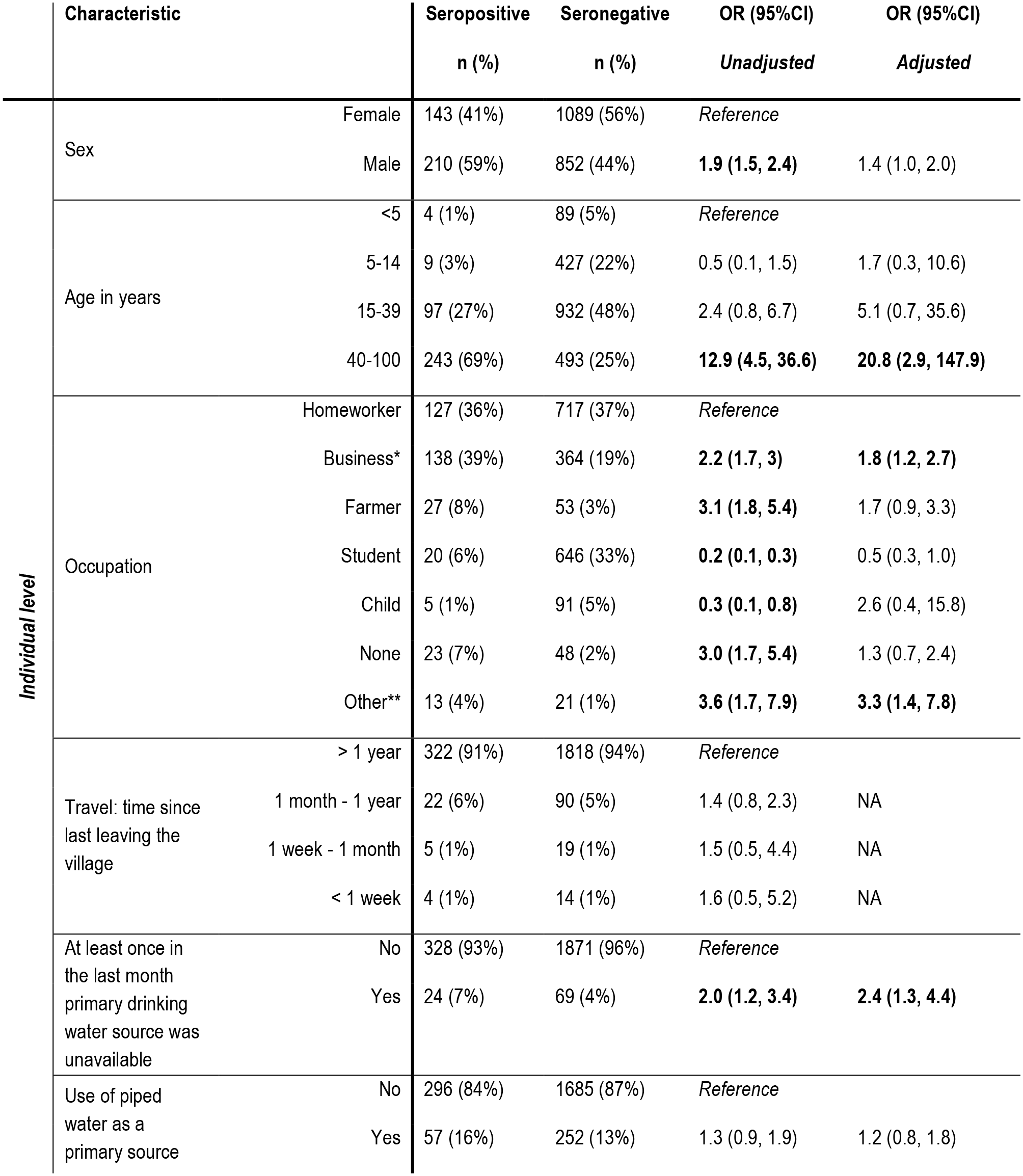
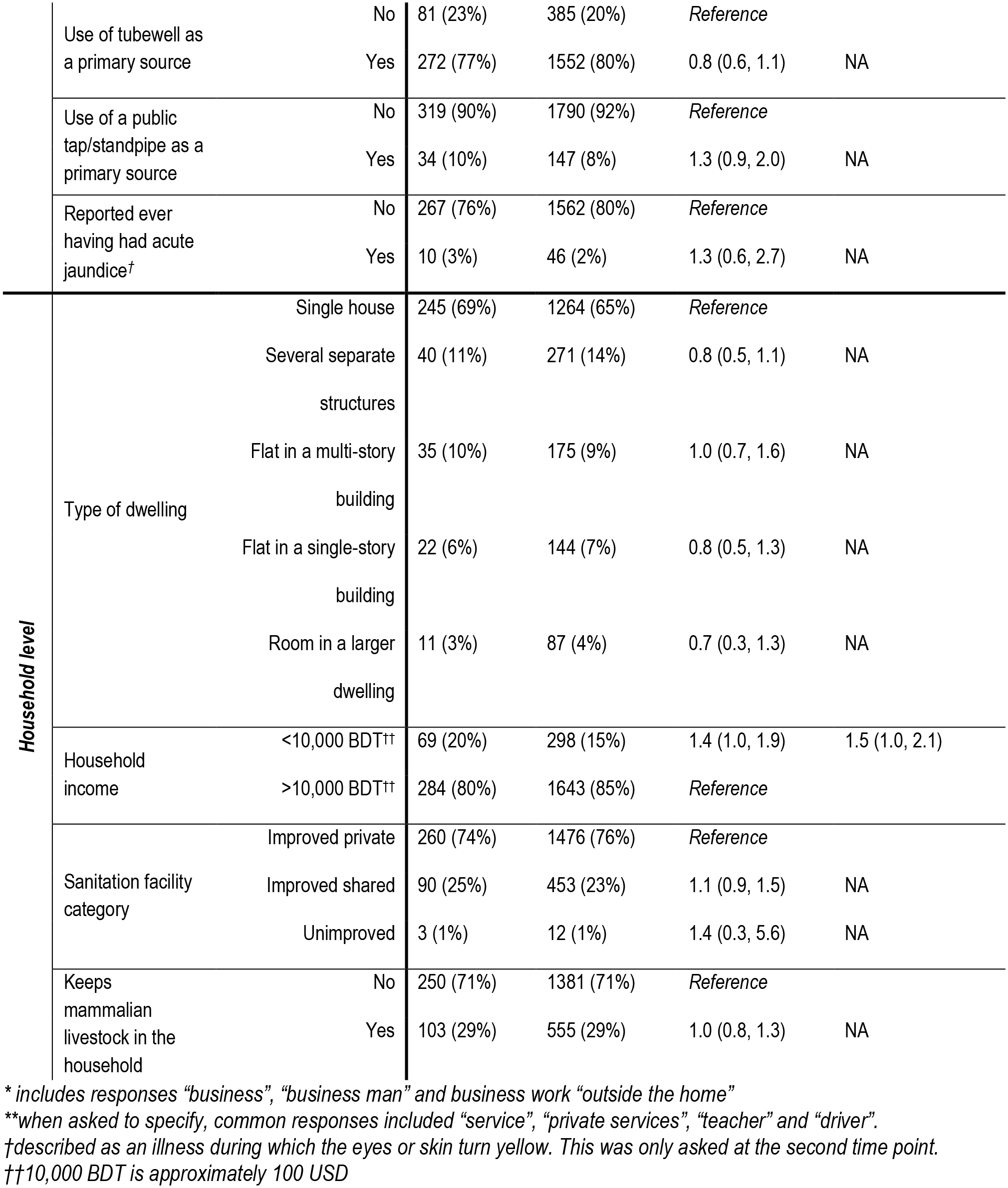
Potential risk factors for past HEV infection at baseline.

Self-reported history of jaundice was not associated with significant increased odds of seropositivity. Of the 59 people who reported having ever had acute jaundice (lasting <3 months), nine reported having had jaundice between survey rounds but none seroconverted. Keeping mammalian livestock was not associated with significantly different baseline seropositivity. Only one household kept pigs – a known host of zoonotic HEV genotypes 3 and 4.

### Evidence of infection during the study period – empirical estimation of the annual risk of infection

Of the 1580 individuals who were seronegative at baseline and provided blood at the end of the study, 38 became seropositive, with similar rates of seroconversion in men and women. This translates to an annual risk of infection of 3% (95% CI: 2-5%) for a seronegative individual in Sitakunda, or approximately 12500 infections in those 1-year-old in 2021. The mean annual risk of infection was higher in adults (4%; 95 CI: 3-6%) than in children <18 years old (3%; 95%CI: 1-5%) but this was not statistically significant. Of the 38 seroconverters, seven lived within the high seroprevalence cluster where the annual infection risk was 19% (95%CI: 7-38%). The 38 seroconverters came from 29 households, and among these, five households had >1 seroconverter, including one where five members became seropositive.

### Waning of antibodies during the study period – empirical estimation of the rate of seroreversion

Of the 266 individuals who were seropositive at baseline and for whom we have paired samples, 27 became seronegative during the study. This translates to an annual risk of seroreversion of 15% (95%CI: 10-21%). Seroreversion rates were slightly lower in males (12%, 95%CI: 6-20%) than females (19%, 95%CI: 11-32%). Seroreversion rates were significantly higher in children than in adults (Figure S3). Of the five children <10 who were seropositive at baseline, four seroreverted. The mean time to seroreversion for children <10 years was estimated to be 7 months (95%CI: 3-24 months) compared to 8 years (95%CI: 5-12 years) for those ≥10 years.

As changes in seropositivity could result from small fluctuations in antibody concentration or measurement error around the cut-off, we compared the optical density to cut-off ratios for individuals across rounds. The majority of seroreverters and seroconverters had a considerable change in antibody titres (Figure S4A-B). Several individuals classified as seropositive at baseline had a substantial increase in their optical density to cut-off ratio suggesting reinfections (Figure S4.C).

### Estimating the annual risk of infection from cross-sectional data

The annual risk of HEV infection has previously been estimated by fitting catalytic models of seroconversion to age-stratified cross-sectional seroprevalence data, assuming antibodies do not wane. When we fitted a model assuming lifelong antibody persistence (Model 1) to our baseline age-stratified seroprevalence data, our estimated annual risk of infection was 5-times lower than what we had measured based on observed seroconversion events (Figure 2). This same magnitude of difference was seen both when we assumed the risk of infection was the same across ages (Model 1a), and when we allowed risk of infection to change around 24-30 years of age (Model 1b, which optimized model fit; Table S3, Figure S5 & S6).

**Figure 2.**
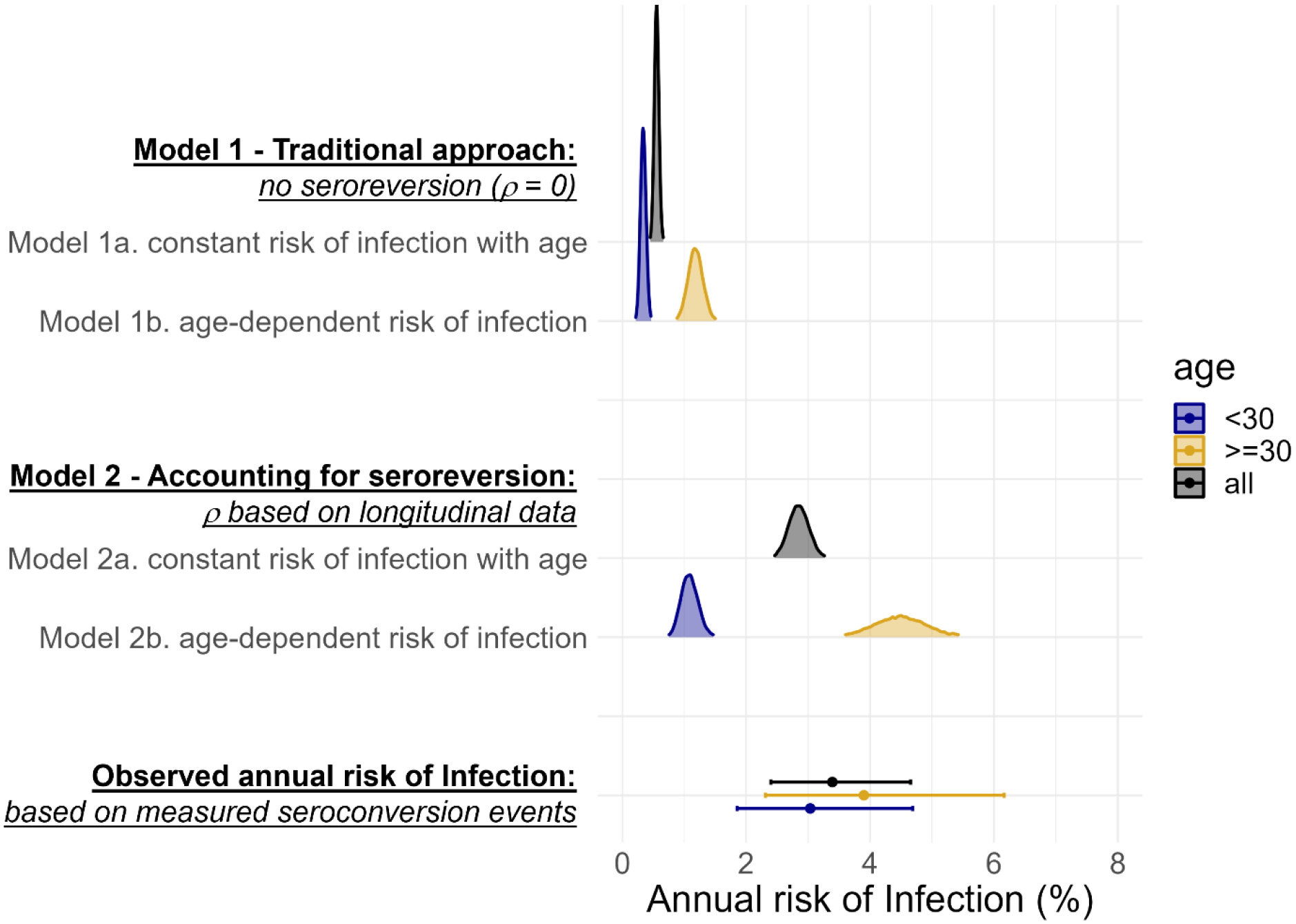
A comparison of posterior estimates of the annual risk of infection from catalytic models 1 and 2 fitted to age-stratified cross-sectional data (smoothed density curves) and the estimates of the annual risk of infection from observed seroconversion events captured in our longitudinal serostatus data (points with bars representing 95%CIs).

When we fitted a model (Model 2) assuming antibodies wane below detectable levels at the rate we measured in the cohort (1.8 (95%CI: 0.5-4.6) in <10-year-olds and 0.1 (95%CI: 0.1-0.2) in ≥10-year-olds), the resulting estimates of annual risk of infection were more comparable to what we measured longitudinally (Figure 2). When we assumed that all age-classes experienced the same risk of infection (Model 2a), the estimated annual risk of infection was 2.8% (95%CrI: 2.5-3.2%) similar to the 3.4% (95%CI: 2.4-4.6%) based on the observed seroconversion events. However, this model fit resulted in overestimates of age-specific seroprevalence for younger age-classes and underestimates for older age-classes when compared to the observed data (Figure S5). Allowing age-class-dependent risk of infection (Model 2b) improved model fit, with the best fit achieved when the risk changed at around 30-years. For individuals >30, the annual risk of infection was 4.5% (95%CrI: 3.7-5.4%), similar to the measured risk for that age group (3.8%, 95%CI: 2.3-6.1%). The annual risk of infection in those <30-years though was lower than what was measured in the cohort (Figure 2). Using the cross-sectional seroprevalence data from follow-up rather than baseline produced very similar estimates of seroprevalence by age and annual risk of infection (Figure S7 & Table S4).

When we attempted to simultaneously fit both the rate of infection and of seroreversion, our estimates were not compatible with the seroreversion rate we measured in the cohort and model fit did not improve (Table S3, Figure S5). When we assumed infection risk was constant with age, the annual risk of seroreversion approached zero (0.09%, 95%CrI: 0.00-0.35%). When we allowed different infection risks for those over and under 30, the annual risk of seroreversion was 5.7% (95%CrI: 2.1-10.7%), less than half what we observed during the study (Figure S8).

## Discussion

In this population-representative longitudinal cohort study, we collected two types of serological data as proxy measures of HEV infection in an endemic region of Bangladesh. We used age-stratified cross-sectional seroprevalence data to explore risk factors associated with past HEV infection and to produce traditional estimates of the annual risk of infection, whilst longitudinal serostatus changes allowed us the opportunity to capture incident infections and seroreversion.

At 15% (95%CI: 14-17%), our estimated baseline seroprevalence was similar to the estimate derived from the 2015 national serosurvey for Sitakunda (19%, 95%CrI: 12-30%) [33]. We estimated that 3% of seronegative people become infected annually in Sitakunda, translating to approximately 12500 HEV infections in people ≥ 1 year old which is approximately half that of previous estimates for rural Bangladesh in 2003-4 (6%, 95%CI: 4-8%) [34]. This could reflect lower transmission intensity in Sitakunda than other rural parts of the country or decreasing incidence over time.

Like previous studies in Bangladesh [33,35], we found male sex to be associated with higher odds of anti-HEV seropositivity in univariate analyses. Our results suggest occupation may confound the effect of sex on infection risk, with those working in business or other occupations outside the household significantly more likely to be seropositive and male. Collecting data on water source use outside the household may help us better understand how these consumption patterns are related to HEV infection risk. Despite evidence that HEV outbreaks have been caused by breeches and contamination of municipal piped water supply systems in urban Bangladesh [36], we did not find use of piped water in the week prior to the survey to be associated with significantly higher odds of seropositivity in this study. However, individuals who reported that their primary water source had been unavailable at least once in the last month were more than 2-times more likely to be seropositive, suggesting that use of water lines with insufficient supply, or having to use alternatives, may increase risk of infection.

In the absence of longitudinal data, estimating annual HEV infection risk has traditionally relied on fitting catalytic models to cross-sectional seroprevalence, assuming life-long antibody persistence and constant risk of infection. By testing paired samples and capturing seroconversion events, we measured the annual risk of HEV infection in Sitakunda to be 5-times higher than the estimate generated by applying traditional approaches to our baseline seroprevalence data. Using observed seroreversion events to inform a reversible model of seroconversion allowed us to obtain estimates of the annual risk of infection closer to what we measured during our study. Simultaneously fitting the rate of seroreversion and infection risk to cross-sectional seroprevalence data could not eliminate the need for empirical measures of seroreversion, likely due to identifiability issues [17]. We found that allowing for different annual infection risk in those >30 greatly improved the model fit, but it was not possible to determine if this reflects a higher risk in those >30 or lower incidence over the past 30 years.

Our estimated annual risk of seroreversion (15%) is higher than existing estimates which were approximately 2% in rural Bangladesh (20% antibody loss over 10-12 years), 4% in Kashmir (50% over 14 years) and 5% in China (30% over 6.5 years) [15,16,29]. We expect this is partly due to the shorter time between paired sample collection in our study compared to previous studies where serorversion events may have been masked by reinfection. The seroreversion rate was significantly higher in children <10 than in adults – a trend also observed in the previous study of anti-HEV IgG loss in Bangladesh [16]. HEV antibodies were also measured to wane quickly in a study in children in Egypt, becoming undetectable in a matter of months [28]. In our study, most children who seroreverted started with optical density values near the upper limit of the dynamic range of the assay suggesting that children may experience faster antibody waning than adults, rather than mounting a lower initial antibody response. Potential reasons for this observed difference in seroreversion rate are not well understood though could include differences in the immune response to HEV in young children, or less antibody persistence due to fewer repeat infections in children compared to adults who have been at risk for longer. A larger sample of seropositive children is needed to investigate these differences in antibody persistence further and to understand the implications for estimating risk of infection in this age group.

Our study has several limitations. Firstly, we used a non-quantitative serological assay and did not use reference serum to allow for generalized comparisons to other studies [37]. We relied on the threshold specified by the kit instructions to classify samples as seropositive and seronegative, assuming perfect assay sensitivity and specificity for detecting infections during the study period. Sensitivity and specificity are estimated to be high [38,39] but the use of this threshold is unlikely to produce perfect, generalisable classification. Although the majority of serostatus changes were associated with large changes in antibody titres, a small minority of seroconversion and reversion events involved relatively small changes. Without a gold standard assay for comparison, we cannot rule out that some serostatus changes could be due to noise. We also saw several large boosts in antibody measures in seropositive individuals that likely represent uncounted reinfections.

Secondly, changes in infection risk over time could contribute to the discrepancy between the seroincidence measured longitudinally and our estimates from cross-sectional data. For example, an undetected outbreak could have elevated the risk of infection measured during our study, but very low numbers of people self-reporting jaundice, and similar seroprevalence to other studies suggest this did not occur. Endemic transmission of HEV is not known to be strongly seasonal in Bangladesh [34], but if there is a seasonal component then we could have slightly biased estimates by not spreading equal at-risk person-time across a full year. Spanning 10.5 months, we would not expect the effect to be considerable in our study. Finally, although we were able to estimate the HEV infection rate in Sitakunda, the absence of a serological assay capable of distinguishing between HEV genotypes, with potentially very different clinical consequences, constrains our ability to translate this into estimates of disease burden. To date, genotyped clinical cases in Bangladesh have all been classified as HEV-1 [18,40], but zoonotic HEV likely circulates in pigs in the country [41] and the incidence of spill over is unknown.

Our results provide evidence of endemic circulation of HEV in Sitakunda, with people who work in occupations outside the home and those reporting their primary water sources to be recently unavailable at higher risk of infection. In the face of widespread under-reporting of hepatitis E cases, estimates of infection incidence from serological data, while imperfect, are important for improving our understanding of transmission, risk and burden of hepatitis E. We were able to evaluate estimates produced by traditional approaches against empirical estimates of seroincidence, demonstrating the need to account for the rate of antibody waning and differences in the risk of infection experienced by different age groups, to avoid underestimating incidence of HEV infection. Refining our interpretation of hepatitis E serological data, through both improving analytical methods and collecting new longitudinal data across geographies, will be key to expanding the breadth of our understanding of this vaccine-preventable disease.

## Data Availability

The data and code produced in this work are available here: https://github.com/HopkinsIDD/sitakunda-hev

## Acknowledgements

We thank all the participants in the cohort study for taking part in this research. We thank the field and laboratory staff for their dedicated work.

## Data and code availability

The data and code used in these analyses are available here: https://github.com/HopkinsIDD/sitakunda-hev

