## Supplementary Material for "Annual risk of hepatitis E virus infection and seroreversion: insights from a serological cohort in Sitakunda, Bangladesh"

*†Equal contributions*

### Contents

**Figure S1.** Location of sampled sites.

**Catalytic model solutions**

**Table S1**. Number of individuals and households sampled during the study.

**Figure S2.** Semivariogram of the semivariance in household level seroprevalence.

**Table S2**: Demographic characteristics of study participants.

**Figure S3**. Annual seroconversion and seroreversion rates by age.

**Figure S4**. The changes in the optical density (od) to cut-off ratios for samples taken at baseline and at follow-up

**Table S3.** The difference in expected log predicted density (ELPD) compared to the preferred catalytic model.

**Figure S5.** Predicted seroprevalence by age.

**Figure S6.** The expected log-predictive density (ELPD) for different age thresholds for changing risk of infection

**Figure S7.** A comparison of the age-stratified seroprevalence by round.

**Table S4.** A comparison of annual risk of infection estimates when using age-stratified seroprevalence at baseline versus at follow-up.

**Figure S8.** A comparison of posterior estimates of the annual risk of infection when we fit the rate of seroreversion, $\rho$, simultaneously with the annual risk of infection, to age-stratified cross-sectional seroprevalence data.

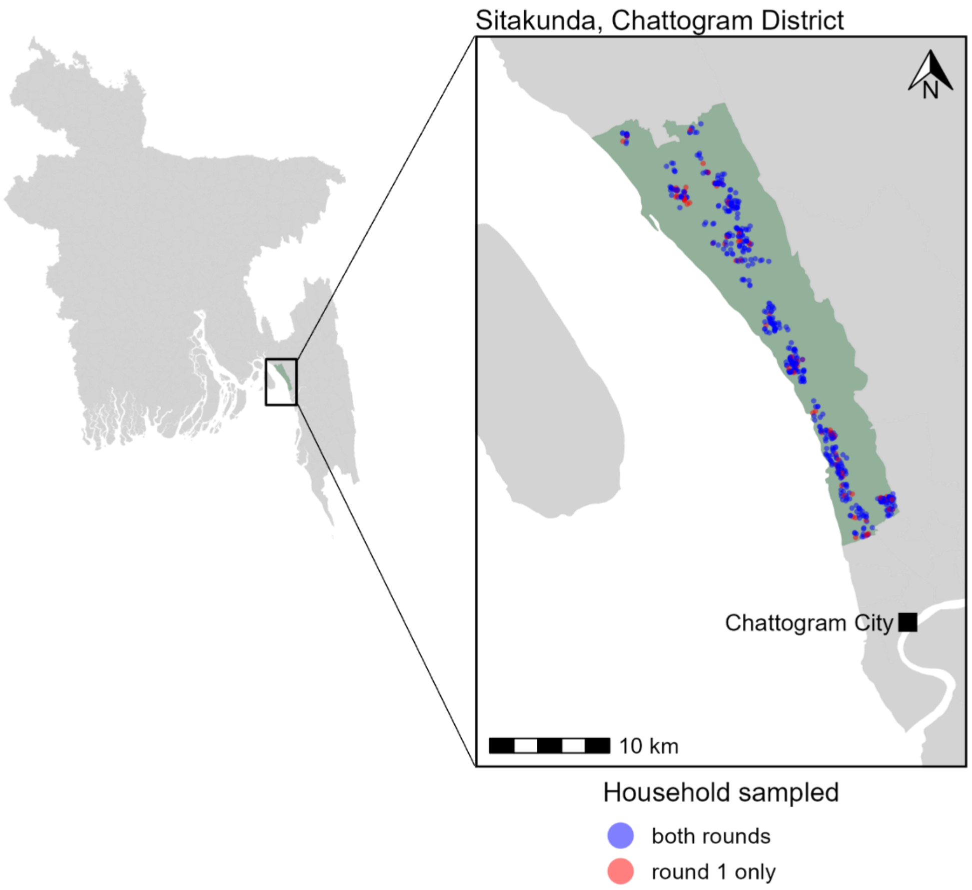


***Figure S1.*** *Location of sampled sites. Sitakunda (green) within Chattogram district, Bangladesh. Sample sites are shown on the inset map, with households sampled in both rounds represented in blue and households sampled at baseline only before being lost to follow-up represented in red.*

### Catalytic model solutions

1) Model 1 - without seroreversion:

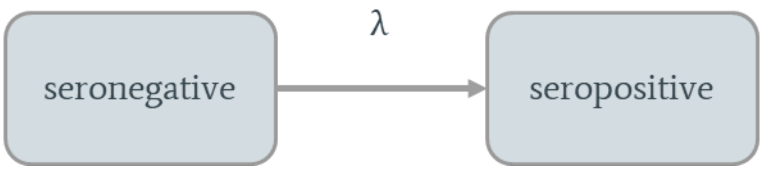


a. Assuming a constant force of infection, λ, with age and time, the proportion seropositive, *p*, at age *a* is:

$$p(a) = 1 - e^{-\lambda a}$$

And so, the proportion seropositive, *p*, in the age class spanning $a_1$-$a_2$ is given by:

$$p(\overline{a_1 \rightarrow a_2}) = \frac{1}{a_2 - a_1}\left(a_2 - a_1 + \frac{1}{\lambda}\left(e^{-\lambda a_2} - e^{-\lambda a_1}\right)\right)$$

b. Assuming the force of infection can be different in children ≤$a_0$ years old and adults >$a_0$

i. When a ≤$a_0$ the proportion seropositive at age a is:

$$p(a) = 1 - e^{-\lambda_C a}$$

And so, the proportion seropositive in the age class spanning $a_1$-$a_2$ where both are ≤$a_0$ is:

$$p(\overline{a_1 \rightarrow a_2}) = \frac{1}{a_2 - a_1}\left(a_2 - a_1 + \frac{1}{\lambda_C}\left(e^{-\lambda_C a_2} - e^{-\lambda_C a_1}\right)\right)$$

ii. For adults, where age a>$a_0$, the proportion seropositive at age a is:

$$p(a) = 1 - e^{-\lambda_C a_0 - \lambda_A (a - a_0)}$$

And so, the proportion seropositive in the age class spanning $a_1$-$a_2$ where both are >$a_0$ is:

$$p(\overline{a_1 \rightarrow a_2}) = \frac{1}{a_2 - a_1}\left(a_2 - a_1 + \frac{1}{\lambda_A}\left(e^{-(\lambda_C a_0 + \lambda_A (a_2 - a_0))} - e^{-(\lambda_C a_0 + \lambda_A (a_1 - a_0))}\right)\right)$$

2) Model 2 - with seroreversion:

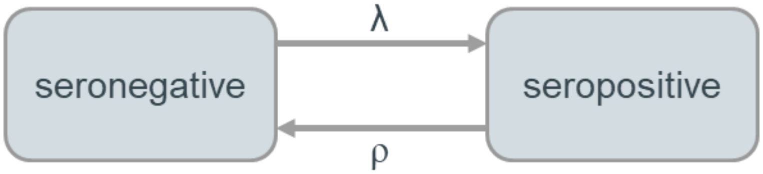


a. Assuming a constant force of infection, $\lambda$, with age and time, the proportion seropositive, *p*, at age *a* is:

$$p(a) = \frac{\lambda}{\lambda+\sigma}\left(1 - e^{-(\lambda+\sigma)a}\right)$$

And so, the proportion seropositive, *p*, in the age class spanning $a_1$-$a_2$ is given below, where $\sigma$ is the annual rate of seroreversion.

$$p(\overline{a_1 \to a_2}) = \frac{1}{a_2 - a_1}\left(\frac{\lambda}{\lambda+\sigma}\left((a_2 - a_1) + \frac{1}{\lambda+\sigma}\left(e^{-(\lambda+\sigma)a_2} - e^{-(\lambda+\sigma)a_1}\right)\right)\right)$$

b. Assuming the force of infection and the annual rate of seroreversion vary with age.
The force of infection changes at age $a_\lambda$ and the annual rate of seroreversion changes at age $a_\sigma$.
We assume $a_\lambda > a_\sigma$.

i. When $a \leq a_\sigma$: the proportion seropositive at age a is:

$$p_1(a) = \frac{\lambda_I}{\lambda_I+\sigma_I}\left(1 - e^{-(\lambda_I+\sigma_I)a}\right)$$

And so, when we solve for the age class spanning $a_1$-$a_2$, where both are $\leq a_\sigma$, we get:

$$p_1(\overline{a_1 \to a_2}) = \frac{1}{a_2 - a_1}\left(\frac{\lambda_I}{\lambda_I+\sigma_I}\left((a_2 - a_1) + \frac{1}{\lambda_I+\sigma_I}\left(e^{-(\lambda_I+\sigma_I)a_2}\right) - e^{-(\lambda_I+\sigma_I)a_1}\right)\right)\right)$$

ii. When $a_\sigma < a \leq a_\lambda$

$$p_2(a) = 1 - \left[\left(1 - p_1(a_\sigma)\right) * \left(1 - \frac{\lambda_I}{\lambda_I+\sigma_{II}}\left(1 - e^{-(\lambda_I+\sigma_{II})(a-a_\sigma)}\right)\right)\right]$$

And so, when we solve for the age class $a_1$-$a_2$, where both are $>a_\sigma$ and $\leq a_\lambda$, we get:

$$p_2(\overline{a_1 \rightarrow a_2}) = \frac{1}{a_2 - a_1}\Bigg[(a_2 - a_1) - \big(1 - p_1(a_\sigma)\big) * \left((a_2 - a_1) * \left(1 - \frac{\lambda_I}{\lambda_I + \sigma_{II}}\right) - \frac{1}{\lambda_I + \sigma_{II}} * \frac{\lambda_I}{\lambda_I + \sigma_{II}} * e^{(\lambda_I + \sigma_{II})a_\sigma} * (e^{-(\lambda_I + \sigma_{II})a_2} - e^{-(\lambda_I + \sigma_{II})a_1})\right)\Bigg]$$

iii. When a> $a_\lambda$

$$p_3(a) = 1 - \left[\big(1 - p_2(a_\lambda)\big) * \left(1 - \frac{\lambda_{II}}{\lambda_{II} + \sigma_{II}}(1 - e^{-(\lambda_{II} + \sigma_{II})(a - a_\lambda)})\right)\right]$$

And so, when we solve for the age class $a_1$-$a_2$, where both are >$a_\sigma$ and ≤ $a_\lambda$, we get:

$$p_3(\overline{a_1 \rightarrow a_2}) = \frac{1}{a_2 - a_1}\Bigg[(a_2 - a_1) - \big(1 - p_2(a_\lambda)\big) * \left((a_2 - a_1) * \left(1 - \frac{\lambda_{II}}{\lambda_{II} + \sigma_{II}}\right) - \frac{1}{\lambda_{II} + \sigma_{II}} * \frac{\lambda_{II}}{\lambda_{II} + \sigma_{II}} * e^{(\lambda_{II} + \sigma_{II})a_\lambda} * (e^{-(\lambda_{II} + \sigma_{II})a_2} - e^{-(\lambda_{II} + \sigma_{II})a_1})\right)\Bigg]$$

***Table S1.*** *Number of individuals and households sampled during the study.*

| | **R1: Mar-Jun 2021** | | **R2: Jan-Feb 2022** | | **Both rounds** | |
|---|---|---|---|---|---|---|
| | individuals | households | individuals | households | individuals | households |
| **Participated** | 2337 | 580 | 2038 | 494 | 1993 | 494 |
| **Blood sampled** | 2308 | 580 | 1986 | 493 | 1862 | 493 |
| **HEV result available** | 2301 | 580 | 1986 | 493 | 1856 | 492 |
| **Excluding borderline results** | 2294 | 579 | 1978 | 492 | 1846 | 491 |

*Note that one household enrolled at round 1 had split into two new households by r2, but this table still counts them as a single household followed up.*

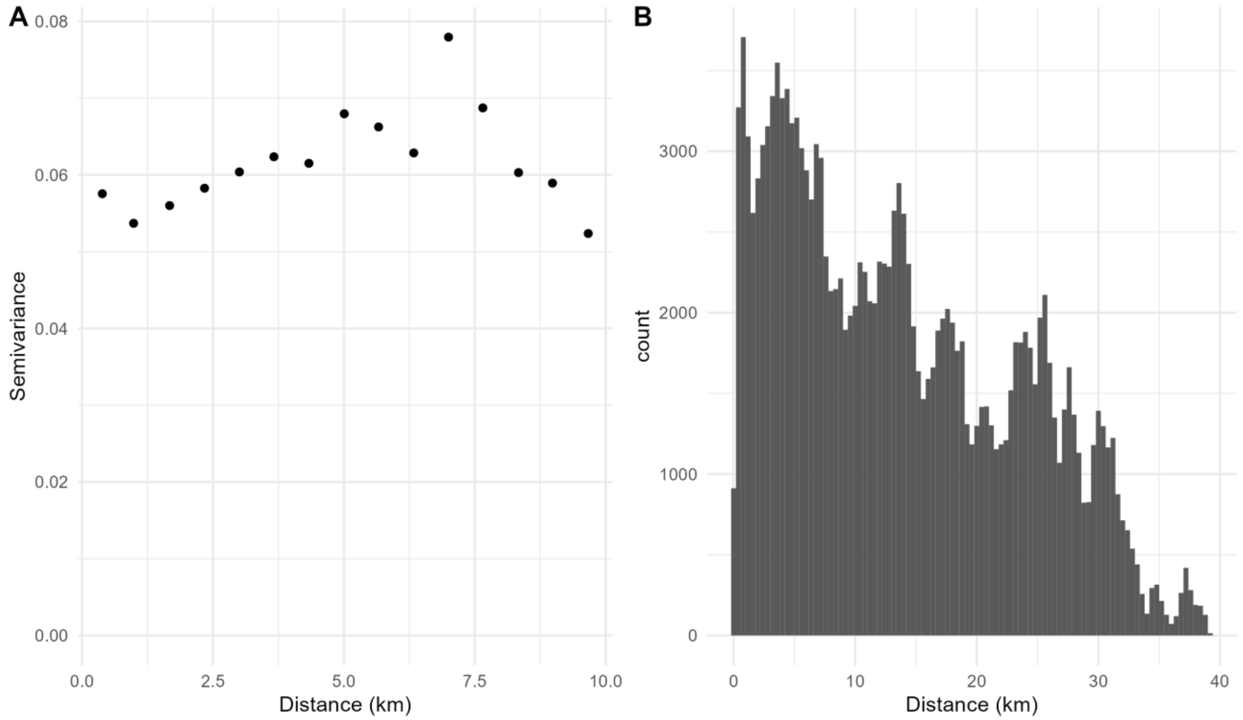


***Figure S2: A****. The semivariogram showing the semivariance in household level seroprevalence plotted against the distance between households.* ***B.*** *The distribution of distance between sampled households.*

***Table S2**: demographic characteristics of study participants, shown overall, split by sex and for the community within the cluster of high seroprevalence in southeast Sitakunda.*

| | Characteristic | | all tested n (%) | male n (%) | female n (%) | cluster* n (%) |
|---|---|---|---|---|---|---|
| Individual level | Sex | Female | 1232 (54%) | NA | 1232 (100%) | 65 (47%) |
| | | Male | 1062 (46%) | 1062 (100%) | NA | 72 (53%) |
| | Age in years | <5 | 93 (4%) | 53 (5%) | 40 (3%) | 6 (4%) |
| | | 5-14 | 436 (19%) | 213 (20%) | 223 (18%) | 23 (17%) |
| | | 15-39 | 1029 (45%) | 444 (42%) | 585 (47%) | 73 (53%) |
| | | 40-100 | 736 (32%) | 352 (33%) | 384 (31%) | 35 (26%) |
| | Occupation | Homeworker | 844 (37%) | 78 (7%) | 766 (62%) | 47 (34%) |
| | | Business* | 502 (22%) | 438 (41%) | 64 (5%) | 42 (31%) |
| | | Farmer | 80 (3%) | 79 (7%) | 1 (0%) | 6 (4%) |
| | | Student | 666 (29%) | 345 (32%) | 321 (26%) | 30 (22%) |
| | | Child | 96 (4%) | 51 (5%) | 45 (4%) | 4 (3%) |
| | | None | 71 (3%) | 41 (4%) | 30 (2%) | 2 (1%) |
| | | Other | 34 (1%) | 29 (3%) | 5 (0%) | 6 (4%) |
| | | NA | 1 (0%) | 1 (0%) | NA | NA |
| | Travel: time since last leaving the village | > 1 year | 2140 (93%) | 982 (92%) | 1158 (94%) | 129 (94%) |
| | | 1 month - 1 year | 112 (5%) | 59 (6%) | 53 (4%) | 5 (4%) |
| | | 1 week - 1 month | 24 (1%) | 10 (1%) | 14 (1%) | 3 (2%) |
| | | < 1 week | 18 (1%) | 11 (1%) | 7 (1%) | NA |
| | Primary drinking water source was unavailable at least once in the past month | No | 2199 (96%) | 1019 (96%) | 1180 (96%) | 108 (79%) |
| | | Yes | 93 (4%) | 41 (4%) | 52 (4%) | 28 (20%) |
| | | NA | 2 (0%) | 2 (0%) | NA | 1 (1%) |
| | Water source category | improved | 2287 (100%) | 1058 (100%) | 1229 (100%) | 136 (99%) |
| | | unimproved | 3 (0%) | 1 (0%) | 2 (0%) | NA |
| | | NA | 4 (0%) | 3 (0%) | 1 (0%) | 1 (1%) |
| | Use of piped water in the past week | No | 1981 (86%) | 910 (86%) | 1071 (87%) | 105 (77%) |
| | | Yes | 309 (14%) | 149 (14%) | 160 (13%) | 31 (23%) |
| | | NA | 4 (0%) | 3 (0%) | 1 (0%) | 1 (1%) |
| | Use of tubewell in the past week | No | 466 (20%) | 225 (21%) | 241 (20%) | 40 (29%) |
| | | Yes | 1824 (80%) | 834 (79%) | 990 (80%) | 96 (70%) |

| | | | | | | |
|---|---|---|---|---|---|---|
| | | NA | 4 (0%) | 3 (0%) | 1 (0%) | 1 (1%) |
| | Use of water from a public tap/standpipe in the past week | No | 2109 (92%) | 971 (91%) | 1138 (92%) | 128 (93%) |
| | | Yes | 181 (8%) | 88 (8%) | 93 (8%) | 8 (6%) |
| | | NA | 4 (0%) | 3 (0%) | 1 (0%) | 1 (1%) |
| | Self-reported ever having had acute jaundice | No | 1829 (80%) | 842 (79%) | 987 (80%) | 105 (77%) |
| | | Yes | 56 (2%) | 33 (3%) | 23 (2%) | 1 (1%) |
| | | NA | 409 (18%) | 187 (18%) | 222 (18%) | 31 (23%) |
| Household level | Type of dwelling | Single house | 1509 (66%) | 712 (67%) | 797 (65%) | 123 (90%) |
| | | Several separate structures | 311 (14%) | 137 (13%) | 174 (14%) | 14 (10%) |
| | | Flat in a multi-story building | 210 (9%) | 94 (9%) | 116 (9%) | NA |
| | | Flat in a single-story building | 166 (7%) | 74 (7%) | 92 (7%) | NA |
| | | Room in a larger dwelling | 98 (4%) | 45 (4%) | 53 (4%) | NA |
| | Household income | <10,000 BDT | 367 (16%) | 166 (16%) | 201 (16%) | 31 (24%) |
| | | >10,000 BDT | 1927 (84%) | 896 (84%) | 1031 (84%) | 105 (76%) |
| | Sanitation facility category | Improved private | 1736 (76%) | 800 (75%) | 936 (76%) | 87 (64%) |
| | | Improved shared | 543 (24%) | 255 (24%) | 288 (23%) | 50 (36%) |
| | | Unimproved | 15 (1%) | 7 (1%) | 8 (1%) | NA |
| | Keeps mammalian livestock in the household | No | 1631 (71%) | 756 (71%) | 875 (71%) | 74 (54%) |
| | | Yes | 658 (29%) | 305 (29%) | 353 (29%) | 63 (46%) |
| | | NA | 5 (0%) | 1 (0%) | 4 (0%) | NA |

**the subset of households located in the high seroprevalence cluster in south-east Sitakunda*

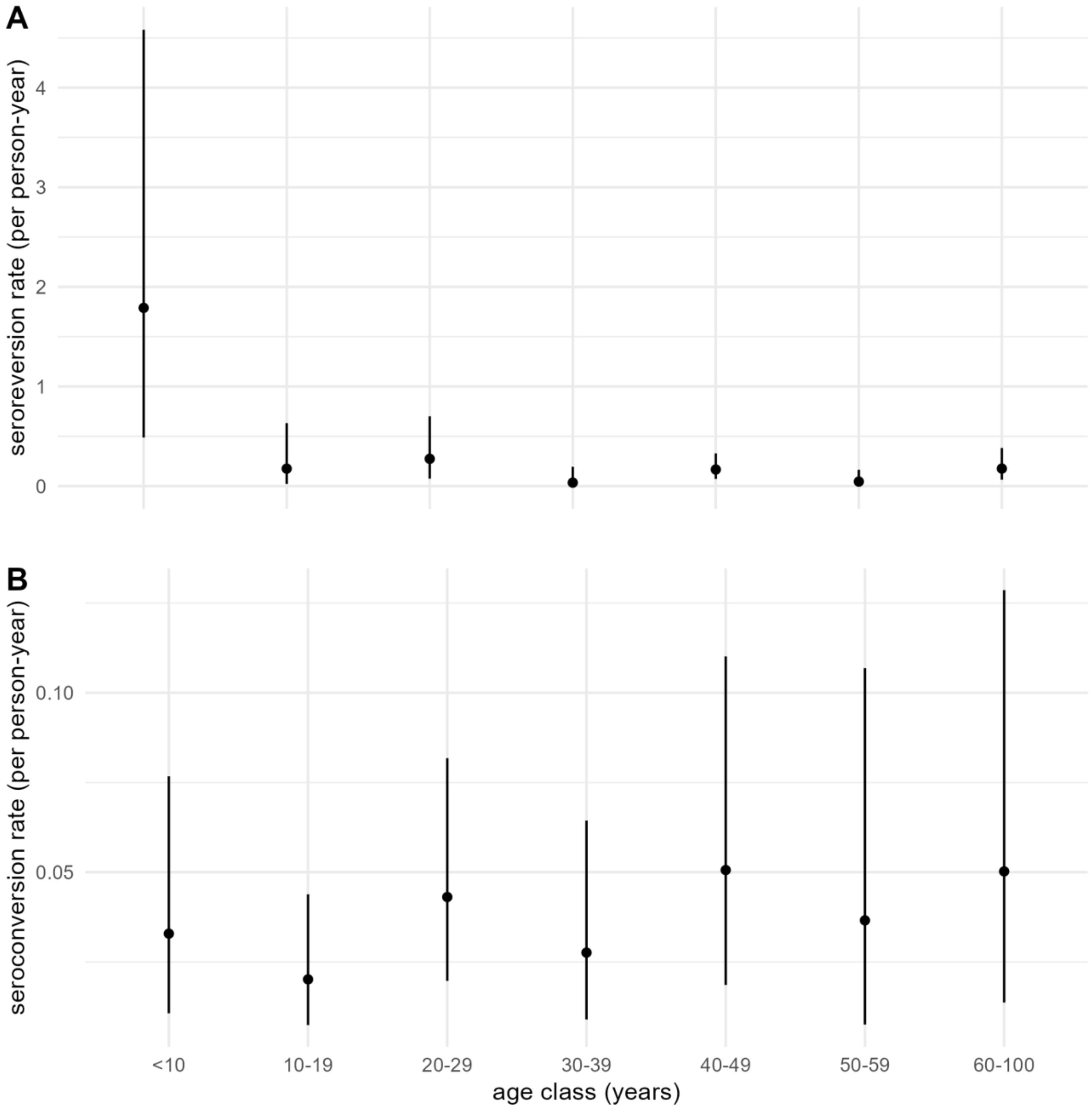


***Figure S3.*** *Annual seroconversion and seroreversion rates by age.* ***A.*** *the annual seroconversion rate per capita by age shown with 95%CIs, accounting for household sampling.* ***B.*** *The annual seroreversion rate per capita by age shown with 95%CIs, accounting for household sampling.*

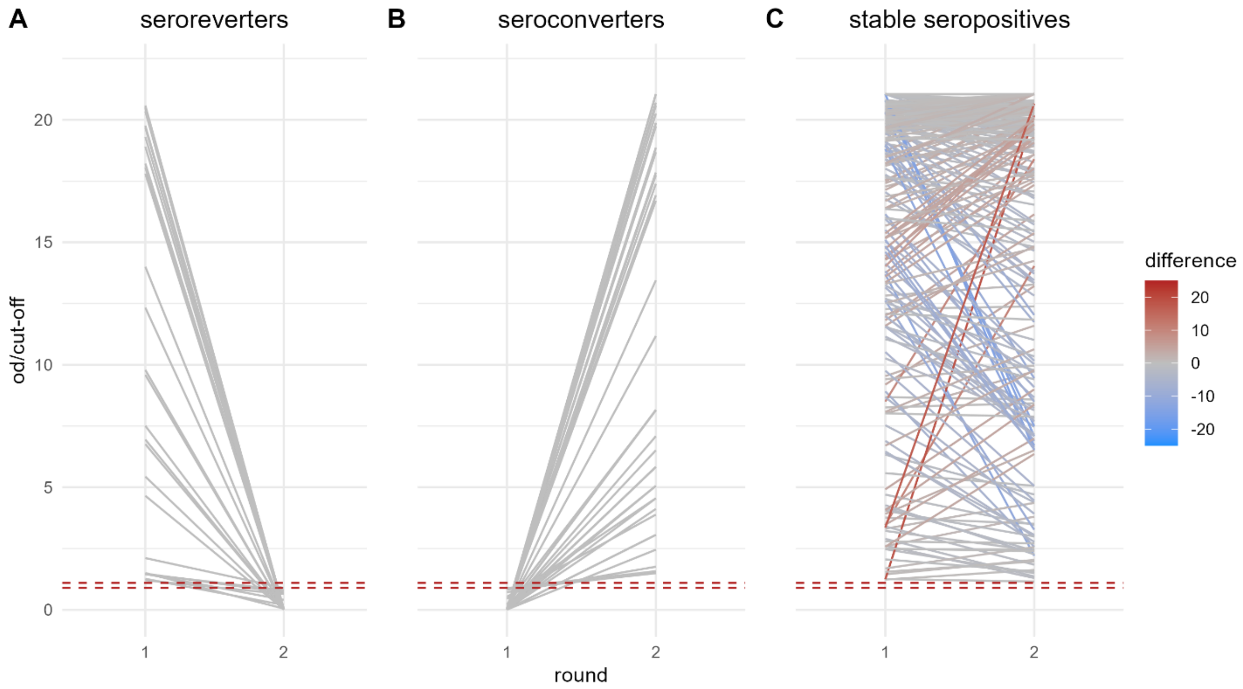


***Figure S4**. The changes in the optical density (od) to cut-off ratios for samples taken at baseline and at follow-up from **A.** individuals who seroreverted during the study, **B.** individuals who seroconverted during the study, and **C**. Individuals who were classed as seropositive at both time points. The area below dashed red lines shows values classed as seronegative, and the area above dashed red lines shows values classed as seropositive, whilst the values between the dashed red lines are borderline and have been excluded.*

***Table S3.*** *The difference in expected log predicted density (ELPD) compared to the preferred catalytic model of seroconversion, as estimated using Leave One Out Cross-validation*

| ELPD difference (SE) | | |
|---|---|---|
| *a. Constant FoI* | b. Age-varying FoI | |
| -22.3 (9.5) | -3.2 (6.6) | Model 1<br>No seroreversion<br>($\rho$ = 0) |
| -72.4 (0.5) | 0.0 (0.0) | Model 2<br>$\rho$ fixed based on longitudinal data |
| -23.4 (9.6) | -1.7 (5.2) | Model 3<br>$\rho$ fitted to cross-sectional data |

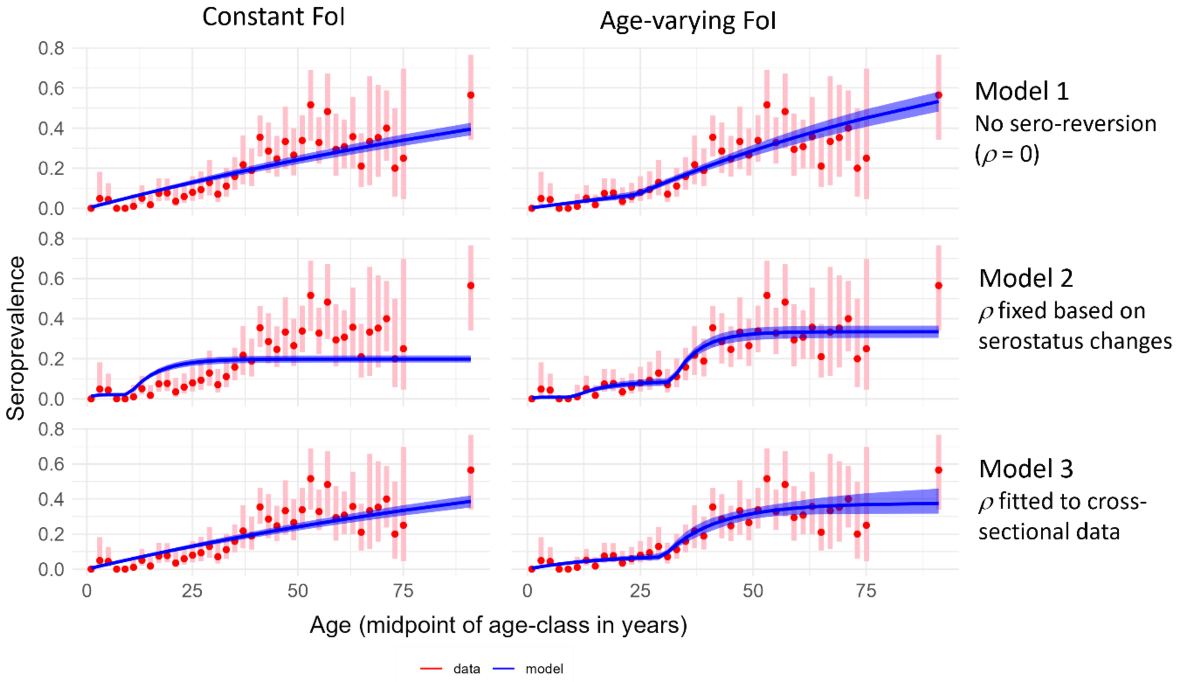


***Figure S5.*** *Predicted seroprevalence by age. Predicted seroprevalence by age is shown in blue for each model, overlaid on the age-stratified baseline seroprevalence measured in our dataset and plotted at the mid-point of each age class. Blue transparent ribbon shows 95%Credible intervals and pink error bars show 95%Confidence intervals for the data.*

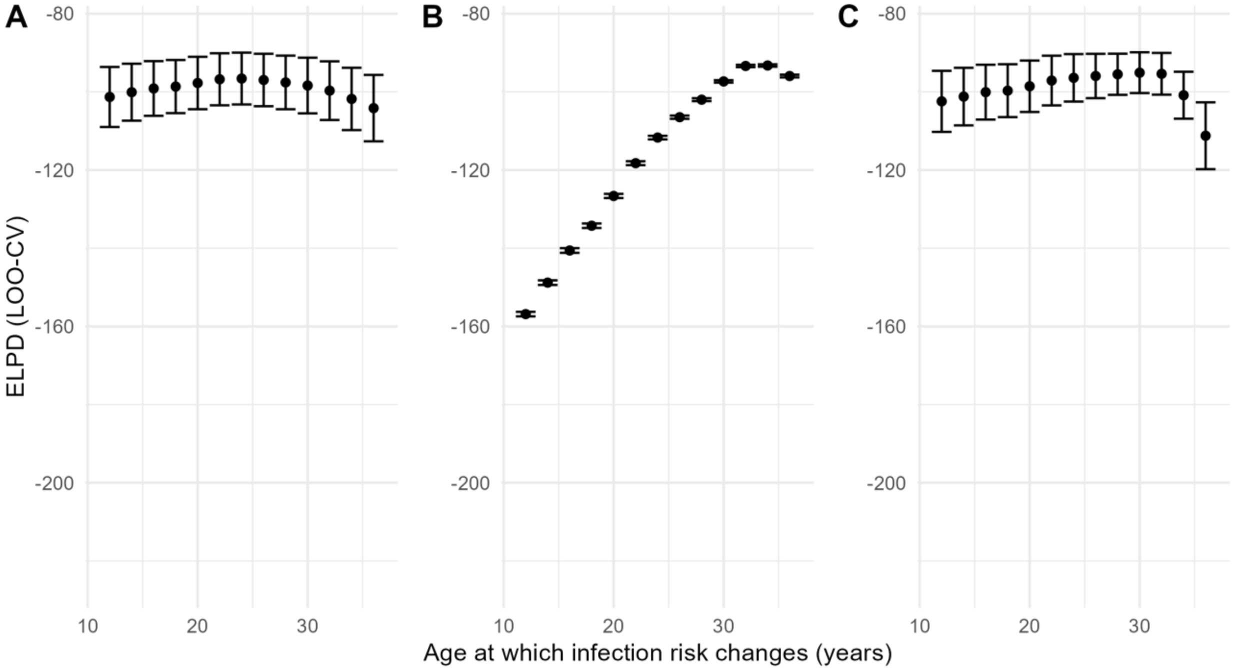


***Figure S6.*** *The expected log-predictive density (ELPD) for different age thresholds for changing risk of infection. The EPLD as estimated using LOO-CV is shown with standard error bars for* ***A.*** *Model 1 in which we assume antibodies last for life (no seroreversion),* ***B.*** *Model 2 assuming instead that the rate of seroreversion that we measured empirically in our cohort, and* ***C****. Model 3 in which the annual risk of infection and the rate of seroreversion are both simultaneously fitted to cross-sectional seroprevalence data.*

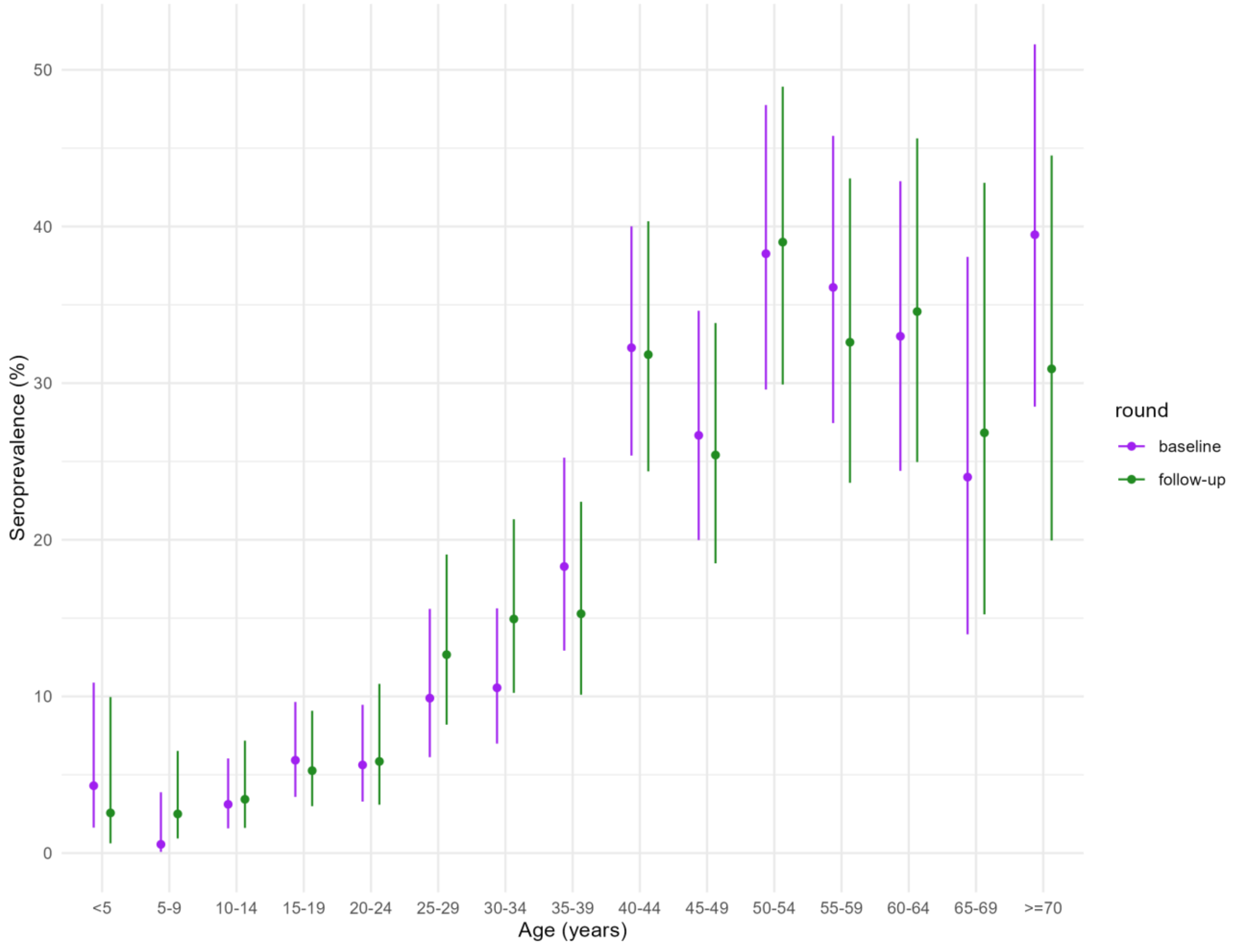


***Figure S7.*** *A comparison of the age-stratified seroprevalence by round.*

***Table S4.*** *A comparison of annual risk of infection estimates when using age-stratified seroprevalence at baseline versus at follow-up.*

| Round | Annual risk of infection (mean and 95% CrI) | |
|---|---|---|
| | <30 year olds | >30 year olds |
| Baseline | 1.1% (0.8-1.4%) | 4.5% (3.7-5.4%) |
| Follow-up | 0.9% (0.7, 1.2%) | 4.1% (3.3-5.1%) |

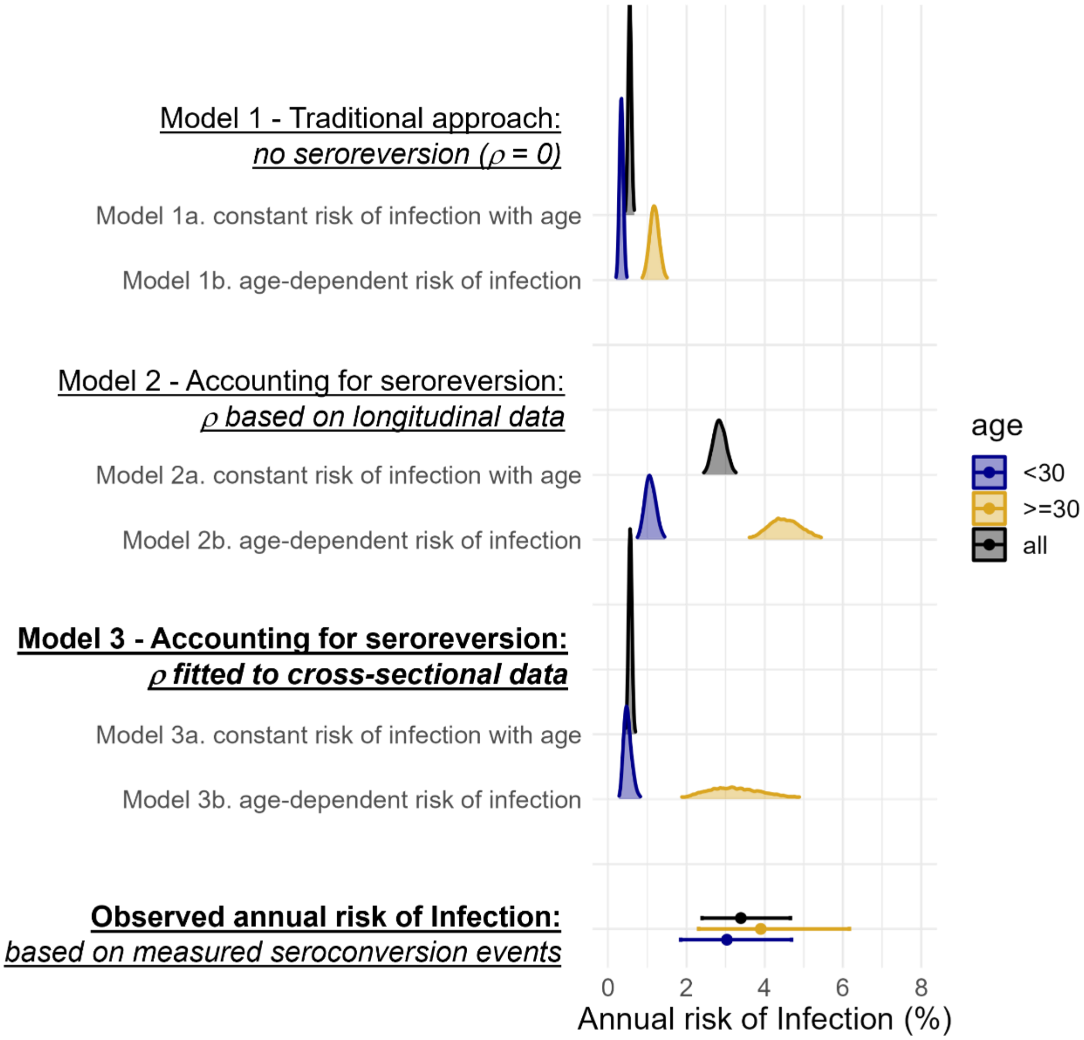


***Figure S8.*** *A comparison of posterior estimates of the annual risk of infection when we fit the rate of seroreversion, $\rho$, simultaneously with the annual risk of infection, to age-stratified cross-sectional seroprevalence data. The results are shown alongside Models 1 and 2 (presented in the main text), along with the estimates of the annual risk of infection from observed seroconversion events captured in our longitudinal serostatus data (points with bars representing 95%CIs.*
